# Levels and determinants of quality of antenatal care in Bangladesh: Evidence from the Bangladesh Demographic and Health Survey

**DOI:** 10.1101/2022.05.31.22275822

**Authors:** Ema Akter, Aniqa Tasnim Hossain, Ahmed Ehsanur Rahman, Anisuddin Ahmed, Tazeen Tahsina, Tania Sultana Tanwi, Nowrin Nusrat, Quamrun Nahar, Shams El Arifeen, Mahbub Elahi Chowdhury

## Abstract

**Background:** Assessing the quality of antenatal care (ANC) is imperative for improving care provisions during pregnancy to ensure the health of mother and baby. In Bangladesh, there is a dearth of research on the quality of ANC using nationally representative data to understand its levels and determinants. The current study aims to assess the quality of ANC and identify the sociodemographic factors associated with the usage of quality ANC services in Bangladesh.

**Methods:** We conducted secondary data analysis using the last two Bangladesh Demographic and Health Surveys (BDHS) (2014 and 2017–18). A total of 8,277 ever-married women were included in the analysis (3,631 from 2014 and 4,646 from 2017–18 BDHS). We constructed the quality ANC index using a principal component analysis on different ANC components: weight, blood pressure measurement, blood and urine tests, counseling about pregnancy complications and a minimum of four ANC visits of which one is by a medically trained provider. Multinomial logistic regression was used to determine the strength of association.

**Results:** Receiving all the six components of quality ANC increased from about 13% in 2014 (BDHS 2014) to 18% in 2017/18 (BDHS 2017–18) with a significant difference of p < 0.001. Women from the poorest group, being rural areas, with no education, high birth order and unexposed to media were less likely to receive high-quality ANC than women from the richest group, from urban areas, with a higher level of education, low birth order and exposure to media.

**Conclusion:** There is a need to improve the quality of ANC services in Bangladesh. An education program for women, with regular knowledge-enhancing sessions for pregnant mothers, may help them understand the value of ANC visits. Documentaries about maternal and child healthcare can be broadcast on television, YouTube, Facebook, radio and other digital platforms regularly.

## Introduction

Assessing the quality of antenatal care (ANC) is imperative for improving care provisions during pregnancy to ensure the health of mother and baby (1). Systematic supervision of a woman during pregnancy is the core intervention within the continuum of care for mothers and babies known as ANC (2). Globally, complications during pregnancy have consistently been a leading cause of maternal death, stillbirth and neonatal death (3). Poor-quality ANC is one contributing factor to this (3). The latest Sustainable Development Goals’ (SDGs) focused on reducing maternal deaths to 70 per 100,000 live births globally by 2030 (4). In addition, the Bangladeshi government has set the goal of reducing the maternal mortality ratio (MMR) from 176 per 100,000 live births to 105 by 2021 (5). It has been established that appropriate quality ANC can save lives (3) and lower maternal mortality by up to 20% (6, 7). Quality ANC has a protective effect against adverse pregnancy outcomes such as low birth weight and premature birth (8-10). In addition, skilled birth attendance (doctors, nurses, and midwives) utilisation during delivery and postnatal care increases with quality ANC (11). Although 64% of pregnant women worldwide attended at least four WHO-recommended ANC activities in 2007–2014 (3), the corresponding figure for Bangladesh was only 47% (BDHS) in 2017–18 (12).

Assessing the quality of ANC necessitates information on service usage of the recommended contents of ANC (13-17). Some literature has defined quality ANC in terms of its required components, which should be delivered during pregnancy for a better, healthier life (3, 12, 18, 19). Recently, the 2016 WHO ANC model has been published, and it recommended a minimum of eight ANC contacts as well as some other components relevant to routine ANC for a successful pregnancy. These include five types of interventions to upgrade the delivery of quality ANC: nutritional interventions, maternal and foetal assessment, preventive measures, interventions for physiological symptoms and health systems interventions (3). A study in Kenya assessed the quality of ANC by evaluating service provisions and the mother’s experience of care, which is an approach based on the WHO model for ANC (3) and Kenya’s National Guidelines for Quality Obstetrics and Perinatal Care, which includes blood pressure measurement, foetal growth exams, urine tests, iron and folic acid supplementation, tetanus toxoid immunisations and at least two doses of intermittent preventive treatment for malaria during pregnancy (18). Physical examination (blood pressure, pulse, temperature) along with minimum basic investigations (haemoglobin, urine, blood group) are considered as appropriate care during ANC according to the Reproductive Maternal Neonatal Child Adolescents Health (RMNCAH) Quality Improvement (QI) Framework (19).

In Bangladesh, large-scale cross-sectional surveys such as BDHS, the Multiple Indicator Cluster Survey (MICS) and the Bangladesh Maternal Mortality and Health Care Survey (BMMS) collect data on ANC by interviewing mothers. These surveys ask women about their weight and blood pressure measurement during ANC, blood and urine samples tests, ultrasonogram test, signs of pregnancy complications, and postpartum family planning. These criteria, along with the criterion of a minimum of four contacts during ANC, have been used to define the concept of quality ANC (12, 20-22).

According to a study conducted in Bangladesh, the technical content of ANC received by mothers during their ANC visits is suboptimal in most facilities, particularly in rural settings (23). Additionally, women are usually unable to receive four or more ANC visits during their pregnancy due to a lack of access to health facilities and skilled health providers (24). An estimated 99% of Bangladeshi health facilities offer ANC services but only 4% are sufficiently prepared to provide quality ANC services in accordance with the WHO guidelines, according to the Bangladesh Health Facility Survey (BHFS) 2017 (25). Appropriate provision of ANC during pregnancy is evidenced by reduced maternal mortalities and afflictions and by healthier motherhoods with better maternal health outcomes (26). Therefore, all pregnant women should have access to maternal health services and it’s beneficial to examine the factors related to these health services reach the goals by minimising pregnancy-related mortality and morbidity (27). A recently published study by Singh et al. (2019) revealed that few studies were conducted to understand the factors that influence ANC quality in low- and middle-income countries (28). Only a few researchers used existing demographic and health survey data to examine the determinants and contents of ANC contacts in Bangladesh (7, 29, 30). The aim of this study is therefore to (a) determine the quality of ANC using the definition mentioned in the BDHS 2017–18 using the last two BDHS surveys (2014 and 2017–18) and (b) examine the factors associated with the usage of the quality of ANC. It is beneficial to conduct this study as there is no study that levels the quality of ANC in Bangladesh using the most updated information.

## Materials and methods

A secondary analysis was conducted by using data from the last two BDHSs (2014 and 2017–18), which are the seventh and eighth in the series undertaken in Bangladesh. They covered all the districts across the administrative divisions of Bangladesh through nationally representative cross-sectional surveys (12, 20). The BDHS is a retrospective study following a multi-stage stratified cluster sampling design. ANC-related information was collected for the last live birth for women who had given birth within three years preceding the BDHS survey. Considering the survey weight, a total of 8,277 ever-married women with their most recent live birth was included in the analysis for the last two BDHSs (2014 and 2017–18), which included 3,631 and 4,646 women, respectively (Fig 1). Information was gathered about the individual, the number of ANC contacts they received and the components delivered in the ANC.

**Fig 1:**
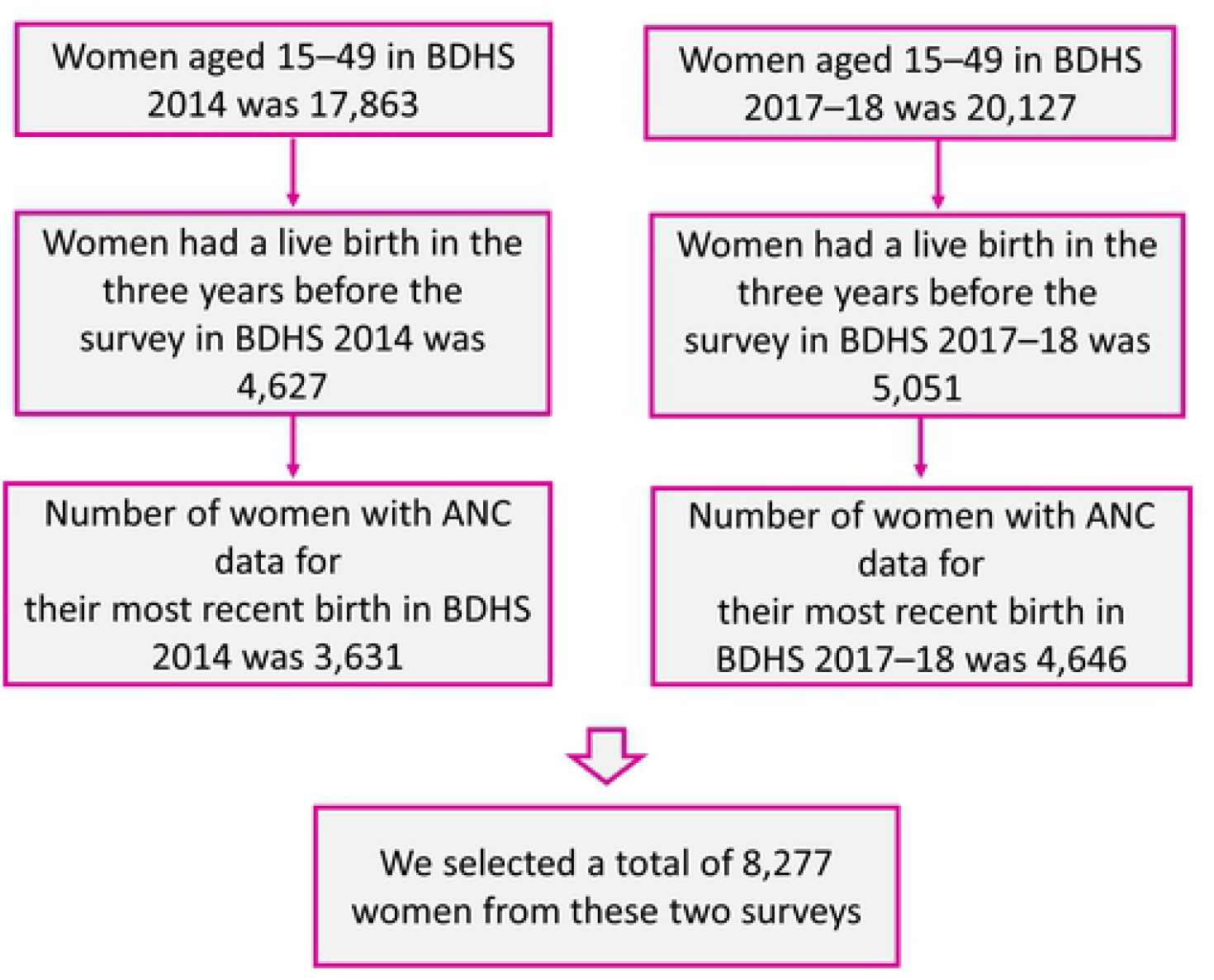
Flowchart of the sample selection process.

This study constructed the quality ANC index using the 2014 and 2017–18 BDHSs, following the definition of quality ANC given in the BDHS 2017–18. The latest BDHS (2017–18) defined quality ANC by considering these components: weight measurement, blood pressure measurement, blood and urine tests, medical advice regarding signs of pregnancy complications and number of ANC visits. For ANC to be considered quality, four visits must take place, one of which must be from a medically trained provider (MTP). We constructed the quality ANCindex based on the above-mentioned components using principal component analysis (PCA). After that, we considered the first principle component as the index obtained from the PCA. We divided this into three categories (low, middle and high). This categorical variable was the outcome variable used in this study, i.e., the quality of ANC tertile.

Several socioeconomic and demographic characteristics were considered as explanatory variables. These included division (Dhaka, Barishal, Chattogram, Khulna, Rajshahi, Rangpur, Sylhet and Mymensingh), place of residence (urban or rural), wealth quintile (the five categories – richest, richer, middle, poorer and poorest – were derived using PCA based on ownership of household goods), mother’s education level (higher, primary, secondary and no education), maternal age in years (15–19, 20–24, 25–29, 30–34 and 35–49), birth order (1, 2–3, 4–5 and 6+), religion (Islam or others) and media exposure (exposed or unexposed). Respondents were considered exposed to media if they performed any of these three actions at least once a week: reading a newspaper or magazine, watching television or listening to the radio. Otherwise, they were considered unexposed to media.

### Statistical analysis

We used descriptive statistics to report the characteristics of the mothers accessing quality ANC. Also, the *p*-value from a chi-square test was reported for both surveys for observing trends over ordered characteristics of quality ANC, according to socioeconomic and demographic characteristics. A 5% level of significance was considered to indicate a significant trend. Multinomial logistic regression was performed for the pooled data from the 2014 and 2017–18 BDHSs to assess the factors affecting the usage of quality ANC while considering the low category of quality ANC as a reference. We reported a crude and adjusted odds ratio (OR) and a corresponding 95% confidence interval (CI) for the multinomial logistic regression. The statistical package Stata 14 (31) was used to perform the analysis.

## Results

Table 1 shows the background characteristics of the mothers who were included in this analysis. The Dhaka division had the highest percentage of women (31%) participating in the survey, while the Barishal and Mymensingh divisions had the lowest percentages of women (5%). Around 71% of the mothers were from rural areas, and 17% were from the poorest socioeconomic group. In both surveys, more than half of the mothers (51%) had a secondary level of education. The 20–24 year age group accounted for the majority of the maternal age group (35%). A large number of mothers (48%) reported a birth order of 2 to 3. Considering religion, around 92% of women were Muslim. Around 58% of the mothers were exposed to media.

**Table 1:** Distribution of the mothers according to background characteristics, presented as frequency and percentage.

| Background characteristics | Survey year 2014 | Survey year 2017–18 | Total |
| --- | --- | --- | --- |
|  | n (%) | n (%) | n (%) |
| **Division** |  |  |  |
| Dhaka | 1,371 (37.8) | 1,204 (25.9) | 2,575 (31.1) |
| Barisal | 196 (5.4) | 245 (5.3) | 441 (5.3) |
| Chittagong | 754 (20.8) | 974 (21.0) | 1,728 (20.9) |
| Khulna | 327 (9.0) | 446 (9.6) | 773 (9.3) |
| Rajshahi | 350 (9.7) | 556 (12.0) | 906 (11.0) |
| Rangpur | 365 (10.1) | 507 (10.9) | 872 (10.5) |
| Sylhet | 267 (7.4) | 327 (7.1) | 595 (7.2) |
| Mymensingh |  | 388 (8.3) | 388 (4.7) |
| **Place of residence** |  |  |  |
| Urban | 1,081 (29.8) | 2,366 (28.6) | 2,366 (28.6) |
| Rural | 2,549 (70.2) | 3,362 (72.4) | 5,911 (71.4) |
| **Wealth status** |  |  |  |
| Wealthiest | 869 (23.9) | 975 (21.0) | 1,844 (22.3) |
| Wealthy | 856 (23.6) | 977 (21.0) | 1,833 (22.1) |
| Middle class | 717 (19.8) | 908 (19.5) | 1,625 (19.6) |
| Poor | 614 (16.9) | 929 (20.0) | 1,543 (18.6) |
| Poorest | 574 (15.8) | 858 (18.5) | 1,432 (17.3) |
| **Educational level** |  |  |  |
| Higher | 450 (12.4) | 854 (18.4) | 1,304 (15.8) |
| Secondary | 1,910 (52.6) | 2,340 (50.4) | 4,250 (51.4) |
| Primary | 898 (24.7) | 1,219 (26.2) | 2,117 (25.6) |
| No education | 373 (10.3) | 234 (5.0) | 607 (7.3) |
| **Age in years** |  |  |  |
| 15–19 | 784 (21.6) | 856 (18.4) | 1,639 (19.8) |
| 20–24 | 1,225 (33.8) | 1,638 (35.3) | 2,863 (34.6) |
| 25–29 | 927 (25.5) | 1,204 (25.9) | 2,131 (25.7) |
| 30–34 | 486 (13.4) | 693 (14.9) | 1,179 (14.3) |
| 35–49 | 208 (5.7) | 256 (5.5) | 465 (5.6) |
| **Birth order** |  |  |  |
| 1 | 1,563 (43.1) | 1,855 (39.9) | 3,418 (41.3) |
| 2–3 | 1,683 (46.3) | 2,290 (49.3) | 3,972 (48.0) |
| 4–5 | 317 (8.7) | 432 (9.3) | 749 (9.1) |
| 6+ | 69 (1.9) | 70 (1.5) | 138 (1.7) |
| **Religion** |  |  |  |
| Islam | 3,354 (92.4) | 4,258 (91.6) | 7,612 (92.0) |
| Others | 277 (7.6) | 388 (8.4) | 665 (8.0) |
| **Exposed to media** |  |  |  |
| Exposed | 2,121 (58.4) | 2,680 (57.7) | 4,801 (58.0) |
| Unexposed | 1,510 (41.6) | 1,967 (42.3) | 3,476 (42.0) |

Fig 2 presents the percentage of mothers receiving specific antenatal services. The percentage of women receiving all six components of quality ANC increased from about 13% (95% CI: 12%– 15%) in 2014(BDHS 2014) to 18% (95% CI: 16%–19%) in 2017–18; (BDHS 2017–18), with a significant difference for these two surveys of p < 0.001. Among the ANC components, blood pressure measurement was the one most commonly received by mothers, with about 88% of mothers receiving it in 2014 and 93% in 2017–18. The second most received component was weight measurement, around 84% in 2014, which increased to 88% in 2017–18. There was also an increase in the provision of the rest of the ANC components from 2014 to 2017–18, with the exception of counselling services about pregnancy complications.

**Fig 2:**
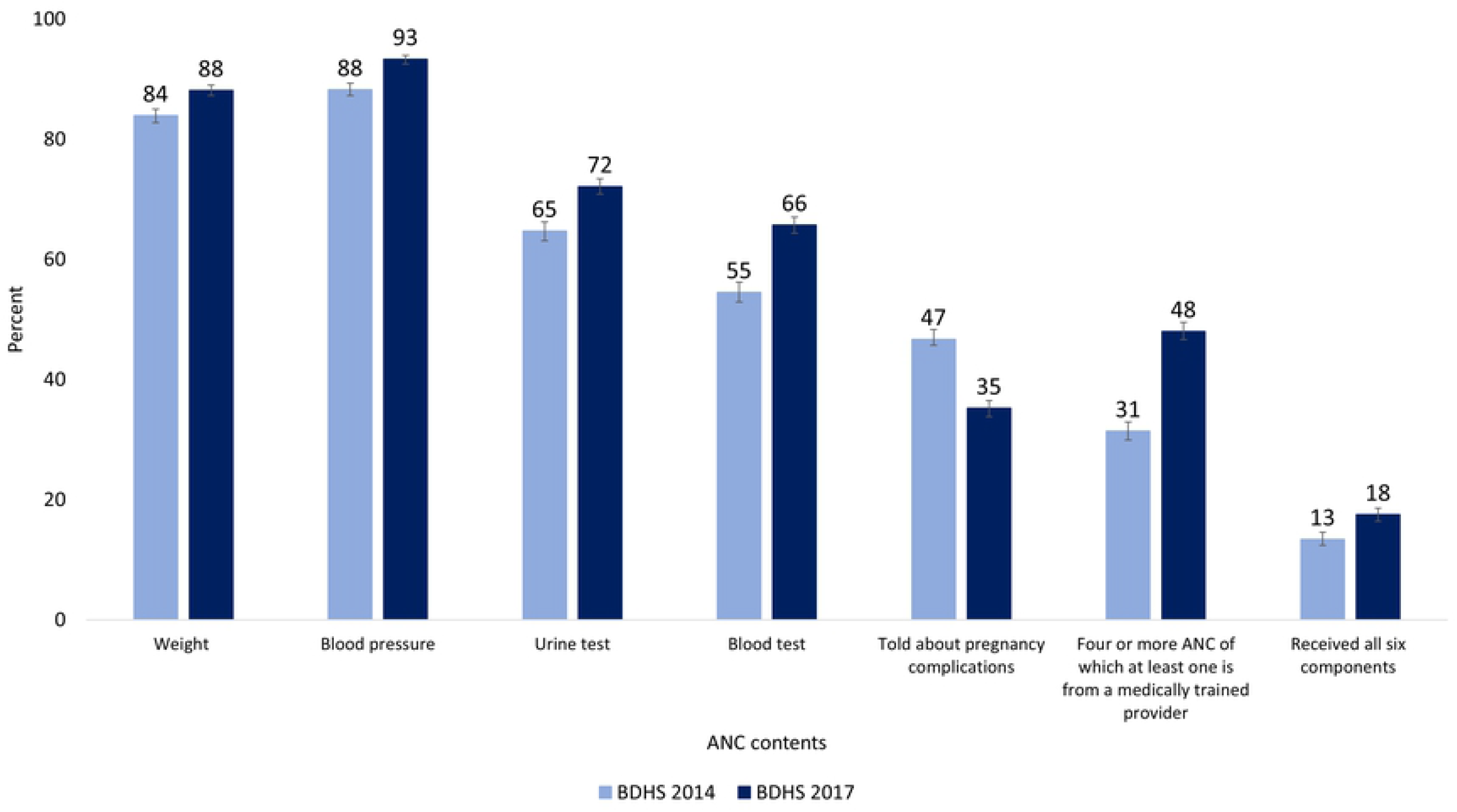
Percentage of mothers receiving specific antenatal services.

Table 2 shows the descriptive statistics regarding the quality of ANC received by background characteristics. The results indicate a significant trend of receiving quality ANC status by type of residence, wealth status, educational level, birth order and media exposure. The percentage of mothers in rural areas receiving high-quality ANC increased by about 15% between the two BDHSs (17% in 2014 and 32% in 2017–18). A large proportion of mothers with no education received low-quality ANC (64% in 2014 and 57% in 2017–18). From 2014 to 2017–18, the percentage of mothers with high quality ANC increased for all age groups and birth orders. A change in trends of receiving quality of ANC status by the mother’s exposure to media status was observed in both surveys; the percentage of mothers exposed to media receiving a high quality of ANC increased by around 15% (31% in 2014 and 46% in 2017–18).

**Table 2:** Descriptive statistics on quality of ANC status by background characteristics, presented as frequency and percentage.

| Background characteristics | Quality of ANC index |  |  |  |  |  |  |  |
| --- | --- | --- | --- | --- | --- | --- | --- | --- |
|  | Survey year 2014 |  |  |  | Survey year 2017–18 |  |  |  |
|  | Low<br>n (%) | Medium<br>n (%) | High<br>n (%) | P-value | Low<br>n (%) | Medium<br>n (%) | High<br>n (%) | P-value |
| **Division** |  |  |  |  |  |  |  |  |
| Dhaka | 605 (44.1) | 410 (29.9) | 356 (26.0) | <0.001 | 353 (29.3) | 304 (25.3) | 547 (45.4) | <0.001 |
| Barisal | 86 (44.0) | 73 (37.4) | 36 (18.6) | <0.001 | 74 (30.2) | 86 (35.3) | 84 (34.5) | 0.337 |
| Chittagong | 296 (39.2) | 292 (38.7) | 167 (22.1) | <0.001 | 325 (33.3) | 322 (33.0) | 328 (33.6) | 0.885 |
| Khulna | 148 (45.2) | 103 (31.4) | 76 (23.3) | <0.001 | 125 (28.1) | 127 (28.6) | 193 (43.3) | <0.001 |
| Rajshahi | 185 (52.8) | 104 (29.8) | 61 (17.5) | <0.001 | 224 (40.2) | 145 (26.0) | 188 (33.8) | 0.022 |
| Rangpur | 183 (50.1) | 116 (31.9) | 65 (18.0) | <0.001 | 200 (39.6) | 132 (26.0) | 174 (34.4) | 0.083 |
| Sylhet | 112 (41.8) | 97 (36.4) | 58 (21.8) | <0.001 | 113 (34.6) | 116 (35.5) | 98 (29.9) | 0.213 |
| Mymensingh | - | - | - | - | 182 (47.1) | 90 (23.3) | 115 (29.6) | <0.001 |
| **Place of residence** |  |  |  |  |  |  |  |  |
| Urban | 358 (33.1) | 329 (30.4) | 395 (36.5) | 0.092 | 299 (23.3) | 322 (25.1) | 663 (51.6) | <0.001 |
| Rural | 1,257 (49.3) | 868 (34.0) | 425 (16.7) | <0.001 | 1,298 (38.6) | 1,000 (29.8) | 1,064 (31.6) | <0.001 |
| **Wealth quintile** |  |  |  |  |  |  |  |  |
| Richest | 180 (20.7) | 280 (32.3) | 409 (47.0) | <0.001 | 113 (11.6) | 214 (22.0) | 648 (66.4) | <0.001 |
| Richer | 321 (37.5) | 312 (36.5) | 223 (26.0) | <0.001 | 253 (25.9) | 306 (31.3) | 418 (42.8) | <0.001 |
| Middle | 385 (53.7) | 229 (32.0) | 103 (14.3) | <0.001 | 349 (38.4) | 263 (29.0) | 296 (32.6) | 0.008 |
| Poorer | 342 (55.6) | 215 (35.0) | 58 (9.4) | <0.001 | 434 (46.8) | 277 (29.8) | 218 (23.4) | <0.001 |
| Poorest | 387 (67.3) | 160 (27.8) | 28 (4.9) | <0.001 | 447 (52.1) | 263 (30.6) | 148 (17.3) | <0.001 |
| **Education level** |  |  |  |  |  |  |  |  |
| Higher | 89 (19.9) | 139 (31.0) | 221 (49.2) | <0.001 | 128 (15.0) | 200 (23.4) | 526 (61.6) | <0.001 |
| Secondary | 792 (41.5) | 664 (34.7) | 454 (23.8) | <0.001 | 762 (32.6) | 662 (28.3) | 916 (39.2) | <0.001 |
| Primary | 496 (55.2) | 298 (33.2) | 104 (11.6) | <0.001 | 574 (47.1) | 394 (32.4) | 250 (20.6) | <0.001 |
| No education | 237 (63.6) | 95 (25.5) | 41 (10.9) | <0.001 | 133 (56.8) | 67 (28.5) | 34 (14.7) | <0.001 |
| **Age in years** |  |  |  |  |  |  |  |  |
| 15–19 | 397 (50.7) | 253 (32.4) | 133 (17.0) | <0.001 | 298 (34.9) | 259 (30.3) | 298 (34.9) | 1.000 |
| 20–24 | 519 (42.4) | 395 (32.3) | 311 (25.4) | <0.001 | 540 (33.0) | 485 (29.6) | 614 (37.5) | 0.006 |
| 25–29 | 402 (43.4) | 305 (33.0) | 219 (23.6) | <0.001 | 397 (33.0) | 332 (27.6) | 475 (39.5) | 0.001 |
| 30–34 | 203 (41.8) | 187 (38.4) | 96 (19.8) | <0.001 | 261 (37.7) | 174 (25.1) | 258 (37.2) | 0.864 |
| 35–49 | 92 (44.2) | 55 (26.5) | 61 (29.4) | 0.001 | 100 (39.0) | 74 (28.8) | 83 (32.3) | 0.111 |
| **Birth rates** |  |  |  |  |  |  |  |  |
| 1 | 638 (40.8) | 509 (32.6) | 416 (26.6) | <0.001 | 525 (28.3) | 532 (28.7) | 797 (43.0) | <0.001 |
| 2–3 | 751 (44.6) | 574 (34.1) | 358 (21.3) | <0.001 | 812 (35.4) | 651 (28.4) | 827 (36.1) | 0.638 |
| 4–5 | 185 (58.5) | 91 (28.8) | 40 (12.7) | <0.001 | 222 (51.5) | 120 (27.8) | 90 (20.8) | <0.001 |
| 6+ | 41 (59.4) | 23 (32.8) | 5 (7.8) | <0.001 | 38 (53.7) | 19 (27.8) | 13 (18.5) | <0.001 |
| **Religion** |  |  |  |  |  |  |  |  |
| Islam | 1,514 (45.1) | 1,098 (32.7) | 742 (22.1) | <0.001 | 1,493 (35.1) | 1,202 (28.2) | 1,564 (36.7) | 0.103 |
| Others | 101 (36.3) | 99 (35.6) | 78 (28.1) | 0.038 | 104 (26.8) | 121 (31.1) | 164 (42.1) | <0.001 |
| **Exposed to media** |  |  |  |  |  |  |  |  |
| Exposed | 751 (35.4) | 706 (33.3) | 664 (31.3) | 0.005 | 698 (26.0) | 738 (27.5) | 1,244 (46.4) | <0.001 |
| Unexposed | 864 (57.2) | 490 (32.5) | 156 (10.3) | <0.001 | 899 (45.7) | 585 (29.7) | 483 (24.6) | <0.001 |

Table 3 shows the results of the multivariate analysis of the socioeconomic and demographic factors associated with mothers receiving quality ANC status. According to the surveys, mothers in 2017–18 had a higher likelihood of receiving high-quality ANC compared with the mothers in 2014. The results showed that place of residence, wealth status, educational level, birth order and media exposure were significant determinants for quality of ANC index. Mothers living in rural areas were less likely to receive high-quality ANC compared with those in urban areas (AOR: 0.78; 95% CI: 0.67–0.90). The adjusted estimate represented higher odd of receiving medium-quality ANC for mothers living in rural areas compared with mothers living in urban areas (AOR: 1.05, 95% CI: 0.91–1.21). Mothers from the poorest group were less likely to receive high-quality ANC compared with mothers from the richest group (AOR: 0.13; 95% CI: 0.10–0.17). Mothers with no education were less likely to receive high-quality ANC compared with mothers with a higher level of education (AOR: 0.20; 95% CI: 0.15–0.28). When compared with older mothers, younger mothers had a lower likelihood of receiving high-quality ANC. Mothers with birth order of six or more had a lower likelihood of having high-quality ANC compared with mothers with the first birth order (AOR: 0.45; 95% CI: 0.25–0.82). Mothers who were not exposed to media were less likely to receive high-quality ANC compared with mothers exposed to media (AOR: 0.65, 95% CI: 0.57–0.75).

**Table 3:** Factors associated with quality of antenatal care received reference category)

| Background characteristics | Middle-quality ANC |  |  | High-quality ANC |  |  |
| --- | --- | --- | --- | --- | --- | --- |
|  | n (%) | Crude OR (95% CI) | AOR (95% CI) | n (%) | Crude OR (95% CI) | AOR (95% CI) |
| **Survey year** |  |  |  |  |  |  |
| 2014 | 1,196 (33.0) | 1.00 | 1.00 | 820 (22.6) | 1.00 | 1.00 |
| 2017–18 | 1,323 (28.5) | 1.12 (1.01–1.24) | 1.23 (1.10–1.37) | 1,727 (37.2) | 2.13 (1.91–2.37) | 2.70 (2.38–3.06) |
| **Division** |  |  |  |  |  |  |
| Dhaka | 715 (27.8) | 1.00 | 1.00 | 903 (35.1) | 1.00 | 1.00 |
| Barisal | 160 (36.2) | 1.34 (1.05–1.70) | 1.63 (1.27–2.08) | 121 (27.4) | 0.80 (0.62–1.03) | 1.37 (1.03–1.83) |
| Chittagong | 613 (35.5) | 1.33 (1.14–1.54) | 1.31 (1.12–1.53) | 494 (28.6) | 0.85 (0.73–0.98) | 0.87 (0.73–1.03) |
| Khulna | 230 (30.0) | 1.13 (0.92–1.38) | 1.17 (0.95–1.45) | 269 (34.9) | 1.05 (0.86–1.27) | 1.25 (1.01–1.56) |
| Rajshahi | 249 (27.5) | 0.82 (0.68–0.98) | 0.87 (0.72–1.05) | 249 (27.5) | 0.65 (0.54–0.78) | 0.79 (0.64–0.98) |
| Rangpur | 248 (28.5) | 0.87 (0.72–1.05) | 1.04 (0.85–1.27) | 240 (27.5) | 0.66 (0.55–0.80) | 1.14 (0.91–1.42) |
| Sylhet | 214 (35.9) | 1.27 (1.03–1.57) | 1.53 (1.23–1.91) | 156 (26.3) | 0.74 (0.59–0.92) | 1.25 (0.96–1.62) |
| Mymensingh | 90 (23.3) | 0.66 (0.51–0.87) | 0.73 (0.55–0.97) | 115 (29.6) | 0.67 (0.52–0.86) | 0.80 (0.60–1.07) |
| **Place of residence** |  |  |  |  |  |  |
| Urban | 651 (27.5) | 1.00 | 1.00 | 1,059 (44.7) | 1.00 | 1.00 |
| Rural | 1,868 (31.6) | 0.74 (0.65–0.84) | 1.05 (0.91–1.21) | 1,489 (25.2) | 0.36 (0.32–0.41) | 0.78 (0.67–0.90) |
| **Wealth quintile** |  |  |  |  |  |  |
| Richest | 495 (26.8) | 1.00 | 1.00 | 1056 (57.3) | 1.00 | 1.00 |
| Richer | 618 (33.7) | 0.64 (0.53–0.77) | 0.70 (0.57–0.84) | 640 (34.9) | 0.31 (0.26–0.37) | 0.41 (0.34–0.49) |
| Middle | 492 (30.3) | 0.40 (0.33–0.48) | 0.44 (0.36–0.54) | 399 (24.5) | 0.15 (0.13–0.18) | 0.22 (0.18–0.27) |
| Poorer | 492 (31.9) | 0.38 (0.31–0.45) | 0.46 (0.37–0.57) | 276 (17.9) | 0.10 (0.08–0.12) | 0.17 (0.13–0.21) |
| Poorest | 422 (29.5) | 0.30 (0.25–0.36) | 0.40 (0.31–0.50) | 176 (12.3) | 0.06 (0.05–0.07) | 0.13 (0.10–0.17) |
| **Education level** |  |  |  |  |  |  |
| Higher | 339 (26.0) | 1.00 | 1.00 | 747 (57.3) | 1.00 | 1.00 |
| Secondary | 1,325<br>(31.2) | 0.55 (0.45–0.66) | 0.72 (0.60–0.88) | 1,370<br>(32.2) | 0.26 (0.22–0.30) | 0.52 (0.43–0.62) |
| Primary | 693 (32.7) | 0.42 (0.34–0.50) | 0.66 (0.53–0.81) | 355 (16.8) | 0.10 (0.08–0.12) | 0.27 (0.21–0.33) |
| No education | 162 (26.7) | 0.28 (0.22–0.36) | 0.48 (0.36–0.64) | 75 (12.4) | 0.06 (0.04–0.08) | 0.20 (0.15–0.28) |
| **Age in years** |  |  |  |  |  |  |
| 15–19 | 513 (31.3) | 0.95 (0.79–1.14) | 0.71 (0.56–0.90) | 431 (26.3) | 0.81 (0.68–0.98) | 0.60 (0.46–0.77) |
| 20–24 | 880 (30.7) | 1.07 (0.91–1.26) | 0.77 (0.56–0.90) | 924 (32.3) | 1.15 (0.97–1.35) | 0.70 (0.56–0.87) |
| 25–29 | 637 (29.9) | 1.03 (0.86–1.22) | 0.87 (0.72–1.05) | 694 (32.6) | 1.14 (0.96–1.35) | 0.92 (0.75–1.13) |
| 30–34 | 360 (30.6) | 1.00 | 1.00 | 354 (30.0) | 1.00 | 1.00 |
| 35–49 | 129 (27.7) | 0.86 (0.67–1.12) | 1.00 (0.76–1.33) | 144 (31.0) | 0.98 (0.76–1.27) | 1.41 (1.03–1.92) |
| **Birth rates** |  |  |  |  |  |  |
| 1 | 1,041<br>(30.5) | 1.00 | 1.00 | 1,213<br>(35.5) | 1.00 | 1.00 |
| 2–3 | 1,225<br>(30.8) | 0.88 (0.78–0.98) | 0.84 (0.72–0.97) | 1,186<br>(29.9) | 0.73 (0.65–0.81) | 0.69 (0.59–0.81) |
| 4–5 | 211 (28.2) | 0.58 (0.48–0.70) | 0.55 (0.43–0.71) | 130 (17.3) | 0.31 (0.25–0.38) | 0.36 (0.27–0.48) |
| 6+ | 42 (30.3) | 0.60 (0.41–0.88) | 0.67 (0.43–1.06) | 18 (13.2) | 0.22 (0.13–0.38) | 0.45 (0.25–0.82) |
| **Religion** |  |  |  |  |  |  |
| Islam | 2,300<br>(30.2) | 1.00 | 1.00 | 2,306<br>(30.3) | 1.00 | 1.00 |
| Others | 219 (33.0) | 1.40 (1.15–1.71) | 1.34 (1.09–1.65) | 241 (36.3) | 1.54 (1.27–1.87) | 1.39 (1.11–1.73) |
| **Exposed to media** |  |  |  |  |  |  |
| Exposed | 1,444<br>(30.1) | 1.00 | 1.00 | 1,909<br>(39.8) | 1.00 | 1.00 |
| Unexposed | 1,075<br>(30.9) | 0.61 (0.55–0.68) | 0.84 (0.74–0.95) | 639 (18.4) | 0.27 (0.25–0.31) | 0.65 (0.56–0.75) |

Fig 3 presents the odds ratio values with with 95% CI for the interaction between the place of residence and wealth quintile. The findings revealed that mothers who lived in urban areas and belonged to the poorest group had a lower probability of receiving high-quality ANC than mothers who lived in urban areas and belonged to the richest group (OR: 0.06, 95% CI: 0.03–0.09). The mothers who lived in rural areas and were from the richest group had insignificantly higher odds of receiving medium-quality ANC than mothers who lived in urban areas and were from the richest group (OR: 1.07, 95% CI: 0.80–1.44).

**Fig 3:**
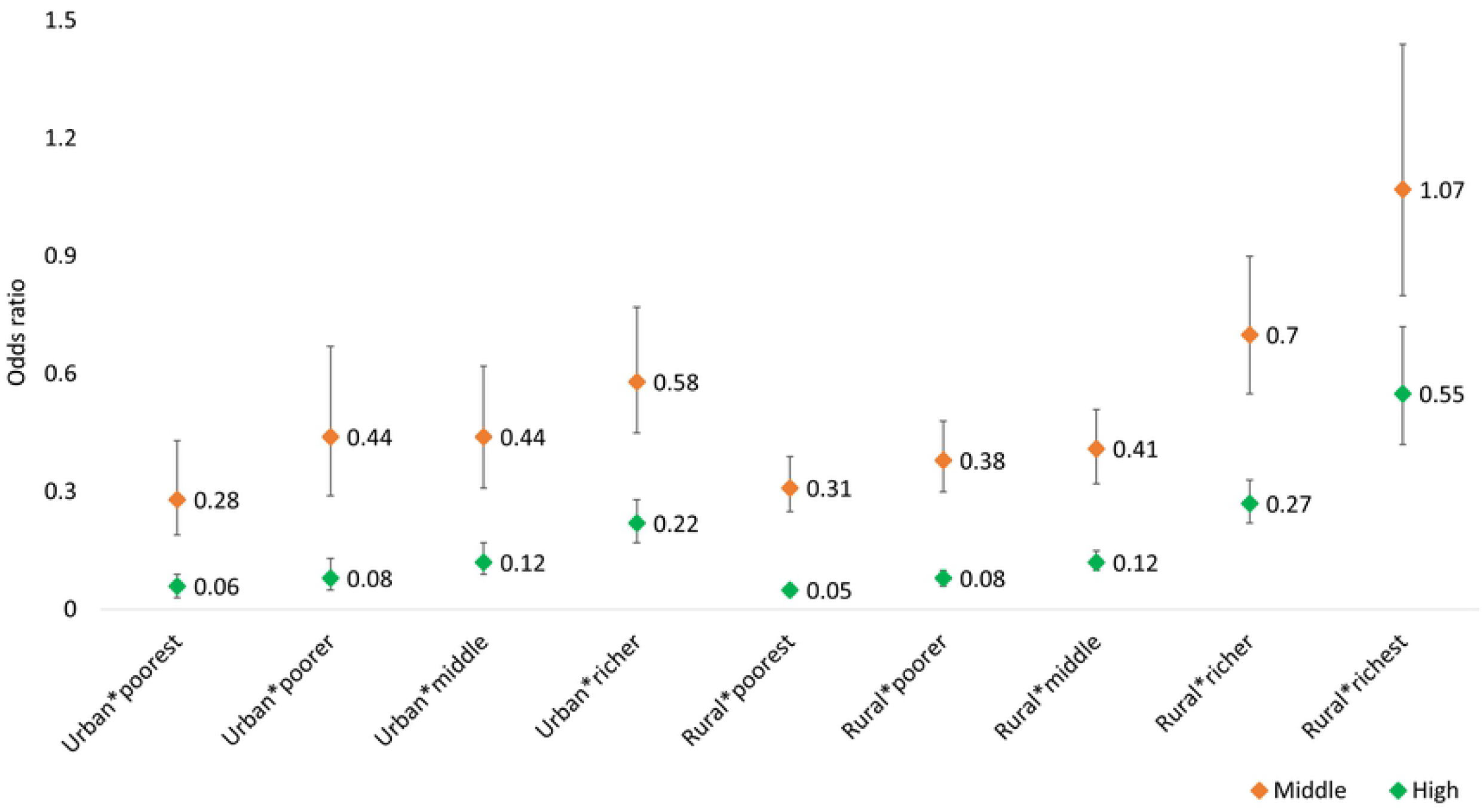
Odds ratio for the interaction between place of residence and wealth quintile (Urban*richest as reference category)

## Discussion

This study reveals that socioeconomic and demographic characteristics are related to receiving better quality ANC and that a considerable proportion of women did not receive adequate ANC with quality. A low percentage of mothers received all six components of quality ANC, with a significant difference between the two surveys (13% in 2014 and 18% in 2017–18 BDHS). Women with high birth order, lower levels of education, belonging to the poorer group, living in a rural area and not exposed to media were less likely to receive high-quality ANC. Around 57% of women from the richest group and around 57% of women with a higher education level received high-quality ANC, whereas only around 12% of women from the poorest group and around 12% of women with no education received high-quality ANC.

This study found that mothers with a higher level of education have a higher likelihood of receiving quality ANC, which is consistent with other studies’ findings from different parts of the world (7, 32-35). A study conducted in Bangladesh documented that the mothers’ educational level strongly affects the optimal uptake of ANC services (7). Uneducated women have less independent decision-making power than educated women (36) and have less knowledge about the importance of using ANC services during pregnancy. Additionally, there are many factors indicating that educated women are more likely to receive quality ANC. Educated women are more conscious about healthcare for themselves and their newborns from the beginning of pregnancy, and as a result, they are willing to access ANC services (37). In addition, women with higher education levels have greater decision-making power about the socioeconomic matters in their households, which give them the confidence and ability to make decisions about their health and the use of quality care services (33, 38).

Differences in place of residence are observed in relation to receiving quality ANC, but interestingly, the adjusted estimate represents a high risk of receiving medium-quality ANC for mothers living in rural areas compared with mothers in urban areas instead of showing a low risk. Interaction is introduced by considering another model, including the place of residence and wealth index in the model. There exists an insignificant interaction between place of residence and wealth. However, this study revealed that women belonging to urban areas have a higher likelihood of receiving high-quality ANC, whereas women belonging to rural areas have a lower likelihood of having high-quality ANC. A study conducted in Nepal had similar findings, documenting that mothers from rural areas are less interested in accessing ANC services than mothers in urban regions (32). Another study in Nigeria reported that un-skilled healthcare providers are factors in rural areas for that women have a lower likelihood of receiving good quality ANC (34). A study in Bangladesh reported that women in urban areas were more likely to receive necessary ANC services than women in rural areas (37). In addition, another study conducted in Bangladesh reported that the lower socioeconomic status of women in rural areas might be associated with a lower uptake of ANC services (7).

A prime determining factor of receiving quality ANC is wealth status. Women with a higher wealth status are more likely to receive a high quality of ANC. This study found that women from the poorest group are less likely to receive high-quality ANC compared with women from the richest group. This finding is consistent with previous studies (7, 28, 34). A study conducted using 91 national household surveys documented that poor women receive a lower quality of care (1). Wealth status could influence the health-seeking nature of women in multiple ways, and without the overall betterment of their standard of living, efforts to encourage poorer women’s utilisation of maternal health services might not obtain useful results (7).

The age of women also affects their uptake of ANC services. The findings from this study show that young women have a lower likelihood of accessing high-quality ANC. Unlike teenage mothers, older women in Bangladesh frequently accessed ANC services (30). A study conducted in Ghana reported that women’s age is associated with the extent of their ANC uptake. Older women are more likely to develop difficulties at birth and have higher uptake of ANC (26). In another study, Tessema & Minyihun (2021) (39) documented that older women understand the benefits of visiting health facilities.

We also found that birth orders are related to uptake of quality ANC services. Women with higher birth order are less likely to utilise high-quality ANC services, implying that women with previous birth experiences demand less ANC. This finding is similar to the results obtained from the study conducted by Nketiah et al. (2013) (26), documenting that in the case of higher births, women are less likely to access ANC than during earlier pregnancies. In addition, a study conducted in Ethiopia reported that women with multiple births are less likely to seek ANC services (2). In Bangladesh, it is apparent that women who give birth for the first time have a higher likelihood of using recommended ANC services (37). This could be due to the belief that they do not need ANC services as they have earlier experiences with pregnancy and childbirth.

Our findings on exposure to media as a factor influencing uptake of ANC services are in line with other studies (7, 37, 40, 41). For instance, a study conducted in Uganda reported that women who have access to media are more likely to use ANC services than women who do not (41). Another study in Nepal found that exposure to media positively impacts the utilisation of ANC components (40). Women exposed to media are more likely to receive ANC services in Bangladesh (37). Women who have access to media are more informed than their counterparts about the usefulness of ANC services during pregnancy and ask the ANC providers for services that may help them lead a healthy life (41). Also, mothers who watch television are more likely to know the problems related to pregnancy and the importance of using ANC services (7).

### Strengths and limitations

We believe this is the first study designing the quality of the ANC tertile using components utilised by women in Bangladesh using nationally representative demographic and health surveys, meaning that its findings are generalisable at the national and sub-national levels. In addition, we have conducted this study using the pooled data of two surveys (2014 and 2017–18 BDHSs), which indicates a larger population-based study for examining the factors associated with the uptake of quality ANC.

The data were collected for three years preceding the survey using the recall method; as a result, there may be some recall bias in the study. However, in the sample, 36.8% of the births were identified for the previous year, 34.8% within 1–2 years and 28.4% within 2–3 years (42). As the recall period varies, the recall bias should be quite low. In the survey, ANC-related information is only available for mothers with live births. Therefore stillbirths and mothers who died during pregnancy or delivery are not included in the study. In addition, we do not have data relating to all of the WHO 2016 ANC model recommendations. However, the BDHS is the best available population based national representative data on the quality of ANC in Bangladesh. We used the BDHS 2017–18 definition of quality ANC as a representation of quality ANC for this study. Due to data limitations, we could not address some important confounding factors, such as cost of care, availability of health facilities and knowledge about maternal healthcare services, that may influence uptake of quality ANC.

## Conclusion and Recommendations

The findings obtained from the analysis provide essential information for policymakers to use to implement the following necessary initiatives to lower maternal and neonatal death by ensuring the quality of maternal health services during pregnancy. Special attention needs to be paid to women in rural areas because they are more deprived than those in urban areas in terms of medical facilities and other amenities. Obstacles to women’s education, such as child marriage and gender-based violence, should be addressed. Re-entry strategies for young mothers should be implemented, just as there should be methods in place to assist women who dropped out of school to return to their studies and the hygiene or sanitation needs of women in schools should be reviewed. In addition, women from early reproductive age groups should be given extra attention. Therefore, there is a need for an education program for women; regular knowledge-enhancing sessions for pregnant mothers may play a vital role in increasing the awareness of the importance of attending ANC visits. The role of media exposure should also be given consideration. Documentaries about maternal health and child healthcare can regularly be broadcast on television, YouTube, Facebook and radio. Drama and Jatra shows could even be staged in rural areas, which would not only will entertain but also will raise awareness. Moreover, trained healthcare providers at the field level should be effectively engaged so that they can go door to door and provide adequate services to pregnant mothers. If the findings and suggestions presented in this paper are acted upon, mothers will become aware of healthcare services during pregnancy.

## Data Availability

The data set can be found from the Demographic and Health Surveys (DHS) program website URL: https://dhsprogram.com/data/available-datasets.cfm

## Acknowledgments

We are grateful to National Institute of Population Research and Training (NIPORT), Bangladesh, and Monitoring and Evaluation to Assess and Use Results Demographic and Health Surveys (MEASURE DHS), USA for allowing permission to utilise the BDHS dataset.

